# Disease burden of bloodstream infections caused by antimicrobial-resistant bacteria: a population-level study—Japan, 2015-2018

**DOI:** 10.1101/2021.02.19.21252053

**Authors:** Shinya Tsuzuki, Nobuaki Matsunaga, Koji Yahara, Keigo Shibayama, Motoyuki Sugai, Norio Ohmagari

## Abstract

**Background:** Antimicrobial resistance (AMR) is a global health problem. However, quantitative evaluation of its disease burden is challenging. This study aimed to estimate the disease burden of bloodstream infections (BSIs) caused by major antimicrobial-resistant bacteria in Japan between 2015 and 2018 in terms of disability-adjusted life-years (DALYs).

**Methods:** We estimated the DALYs of BSIs caused by the six major antimicrobial-resistant bacteria in Japan by utilising comprehensive national surveillance data of all routine bacteriological test results from more than 1,400 hospitals between 2015 and 2018. We modified the methodology of a previous study by Cassini and colleagues to enable comparison of our results with those in other countries.

**Results:** We estimated that 135.8 (95% uncertainty interval [UI] 128.6-142.9) DALYs per 100,000 population was attributable to BSIs caused by the six antimicrobial-resistant bacteria in 2018. *Staphylococcus aureus* (MRSA), fluoroquinolone-resistant *Escherichia coli* (FQREC), and third-generation cephalosporin-resistant *E. coli* (3GREC) accounted for 88.6% of the total. The burden did not decrease during the study period and was highest in people aged 65 years or older.

**Conclusion:** Our results revealed for the first time the disease burden of BSIs caused by six major antimicrobial-resistant bacteria in Japan. The estimated disease burden associated with AMR in Japan is substantial and has not begun to decrease. Notably, the burden from FQREC and 3GREC has increased steadily and that from MRSA is larger than EU/EEA area, whereas that from other bacteria was comparatively small. Our results are expected to provide useful information for healthcare policymakers for prioritising interventions for AMR.

**Funding:** Ministry of Health, Labour and Welfare research grant (20HA2003); Japan Agency for Medical Research and Development Research Program on Emerging and Re-emerging Infectious Diseases (JP19fk0108061)

## Introduction

Antimicrobial resistance (AMR) is a major global health threat (CDC, 2019; European Commission, 2017; World Health Organization, 2015). The World Health Organization published the Global Action Plan on Antimicrobial Resistance in 2015, the strategic objectives of which include improving awareness and understanding of antimicrobial resistance and strengthening knowledge through surveillance and research (World Health Organization, 2015). The following year, the Japanese Ministry of Health, Labour and Welfare published the National Action Plan, which aimed to grasp the state of AMR emergence and its prevalence in Japan (The Government of Japan, 2016).

When considering the implementation of health policies and interventions against AMR, it is necessary to assess its disease burden quantitatively. However, the evidence to date is scarce, especially in the Western Pacific region. Although Tsuzuki and colleagues estimated the number of deaths associated with bloodstream infections (BSIs) caused by methicillin-resistant *Staphylococcus aureus* (MRSA) and fluoroquinolone-resistant *Escherichia coli* (FQREC) in Japan (Tsuzuki et al., 2020), these two organisms represent only a fraction of all antimicrobial-resistant organisms, and “death” is only one aspect of disease burden. Therefore, a more extensive evaluation would be desirable for more precisely understanding the threat of AMR.

Several indicators are used to evaluate disease burden and/or health status, including quality-adjusted life years (QALYs; EuroQol Group, 1990) and disability-adjusted life years (DALYs; Murray and Lopez, 1997). These might be useful indicators for understanding the burden of AMR on our society because they are measurable and comparable.

In the European Union and European Economic Area (EU/EEA), Cassini and colleagues reported the total disease burden of AMR in 2018 in terms of DALYs (Cassini et al., 2018). To our knowledge, their study is the only extensive evaluation of the disease burden of AMR to date. Therefore, we aimed to evaluate the disease burden of AMR in terms of DALYs for the first time in the Western Pacific region and compare our results with theirs.

We focused on BSIs caused by six major antimicrobial-resistant organisms, which were the targets of a comprehensive national surveillance of all routine bacteriological tests performed at more than 1,400 hospitals between 2015 and 2018: MRSA, FQREC, third-generation cephalosporin-resistant *E. coli* (3GREC), third-generation cephalosporin-resistant *Klebsiella pneumoniae* (3GRKP), Carbapenem-resistant *Pseudomonas aeruginosa* (CRPA), and Penicillin-resistant *Streptococcus pneumoniae* (PRSP). Presence of bacteria in a blood specimen almost always means that the patient has a disease; accordingly, disease burden can be estimated from blood culture surveillance data. Additionally, BSIs usually account for the largest part of the burden of infectious diseases due to their high fatality (Anderson et al., 2014; Pittet et al., 1997). Therefore, assessing the disease burden of BSIs would provide a useful indicator for healthcare policymakers.

## Methods

### Data sources

We used data collected by the Japan Nosocomial Infections Surveillance (JANIS) programme organised by the Ministry of Health, Labour and Welfare (Kajihara et al., 2020; Ministry of Health Labour and Welfare Japan, n.d.; Tsutsui and Suzuki, 2018). The JANIS Clinical Laboratory module comprehensively collects all routine microbiological test results, including culture-positive and -negative results from hospitals voluntarily participating in the surveillance, which account for a quarter of the hospitals in Japan.

We extracted the data on MRSA, FQREC, 3GREC, 3GRKP, CRPA, and PRSP isolates from blood specimens collected between 2015 and 2018 in the JANIS Clinical Laboratory database. Patient identifiers were de-identified by each hospital before the data were submitted to JANIS. Approval for extraction and use of the data was granted by the Ministry of Health, Labour and Welfare (0424e1).

Each isolate detected from a blood specimen was counted as one case of BSI. To avoid duplication from the same patient, we included only one specimen from each patient within one year, following the protocol of the European Antimicrobial Resistance Surveillance Network (EARS-Net; Cassini et al., 2018). The judgment criteria for assessing the antimicrobial susceptibility of each bacterium were in accordance with the regulations of JANIS, which follows the criteria defined by the Clinical Laboratory Standards Institute (CLSI; Clinical and Laboratory Standards Institute, 2015).

### Statistical analysis

The number of beds covered by JANIS varies by year and prefecture because the number of participating facilities has increased each year. Therefore, we adjusted the total number of reported BSIs by year and prefecture according to the proportion of the number of beds in participating hospitals, similar to our previous work (Tsuzuki et al., 2020). Each prefecture’s number of beds was calculated as the sum of each participating facility’s number of beds. Information about the total number of beds was obtained from the e-Stat website, a portal site for Japanese Government Statistics (Ministry of Internal Affairs and Communications, n.d.). We excluded psychiatric beds and long-term care beds. Thus, only beds for acute care and infectious diseases were included.

DALYs is a composite health measure estimating both years lived with disabilities (YLDs) following the onset of sequelae and years of life lost (YLLs) due to premature mortality compared with a standard life expectancy (Lier et al., 2007). The number of deaths attributable to the six aforementioned BSIs was estimated by using fatality data obtained from a review of the literature (Cassini et al., 2018; Gallagher et al., 2014; Nagao, 2013; Takeshita et al., 2017), as shown in Supplementary Table 1. To calculate YLLs, we used the estimated number of deaths due to BSIs by age group and life expectancy in Japan. As for YLDs, there is scarce evidence about the incidence of morbidity caused by BSIs in Japan, and thus we used disease model trees and parameters from previous studies conducted in Europe (Colzani et al., 2017; Kretzschmar et al., 2012).

**Table 1.** Estimated number of death due to AMR-BSIs.

|  | 2015 | 2016 | 2017 | 2018 |
| --- | --- | --- | --- | --- |
| **MRSA** | **3834 (2498-5301)** | **3914 (2551-5418)** | **3990 (2601-5510)** | **3902 (2542-5396)** |
| **FQREC** | **3045 (2868-3234)** | **3437 (3239-3649)** | **3561 (3358-3777)** | **3936 (3706-4176)** |
| **3GREC** | **2225 (1197-3266)** | **2321 (1248-3409)** | **2494 (1341-3661)** | **2784 (1497-4090)** |
| **3GRKP** | **483 (362-617)** | **511 (383-653)** | **484 (363-618)** | **421 (561-719)** |
| **CRPA** | **362 (313-413)** | **385 (333-440)** | **310 (268-353)** | **333 (288-380)** |
| **PRSP** | **126 (42-231)** | **108 (36-198)** | **94 (31-173)** | **115 (38-211)** |
| **Total** | **8557 (7178-10000)** | **8902 (7516-10382)** | **9026 (7614-10552)** | **9447 (8068-10910)** |

AMR-BSIs, bloodstream infections caused by antimicrobial-resistant organisms; MRSA,
*Escherichia coli*; 3GREC, third-generation cephalosporin-resistant *E. coli*; 3GRKP,
third-generation cephalosporin-resistant *Klebsiella pneumoniae*; CRPA,
carbapenem-resistant *Pseudomonas aeruginosa*; PRSP, penicillin-resistant

DALYs were calculated using the following equations.

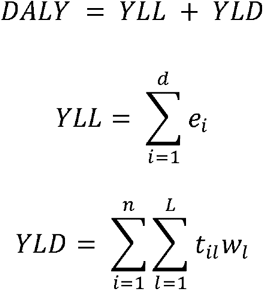

YLLs due to a specific disease in a given population are calculated by summation of all fatal cases (*d*). Each case (*i*) is multiplied by the expected individual lifespan at the age of death (*e*). The YLD is calculated by the product of duration (*t*) and the severity weight (*w*) of a specific health outcome, accumulated over all cases (*n*) and all health outcomes (*L* = 8 ; complicated cases are defined in Supplementary Figure 1). In the present study, all sequelae (i.e. health outcomes except for death derived from complicated BSIs) are assumed to persist throughout life. We assume a time discount rate of 0.02 per year in accordance with the Japanese guidelines (Shiroiwa et al., 2017).

**Figure 1.**
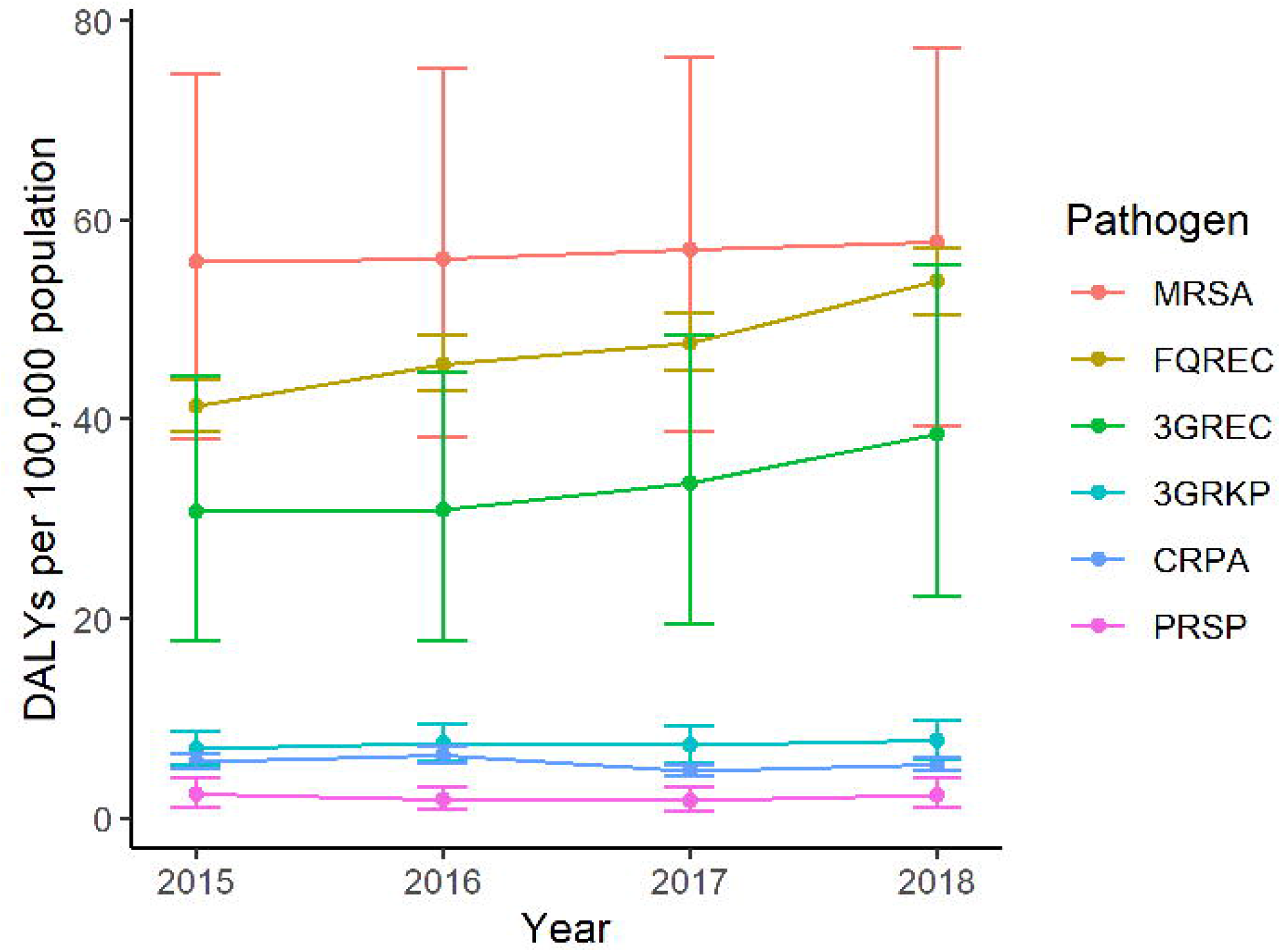
DALYs due to BSIs caused by six major antimicrobial-resistant organisms from 2015 to 2018. DALYs, disability-adjusted life years; BSIs, bloodstream infections; MRSA, methicillin-resistant *Staphylococcus aureus*; FQREC, fluoroquinolone-resistant *Escherichia coli*; 3GREC, third-generation cephalosporin-resistant *Escherichia coli*; 3GRKP, third-generation cephalosporin-resistant *Klebsiella pneumoniae*; CRPA, carbapenem-resistant *Pseudomonas aeruginosa*; PRSP, penicillin-resistant *Streptococcus pneumoniae* Orange line represents DALYs due to BSIs caused by MRSA. Brown line represents DALYs due to BSIs caused by FQREC. Green line represents DALYs due to BSIs caused by 3GREC. Light blue line represents DALYs due to BSIs caused by 3GRKP. Blue line represents DALYs due to BSIs caused by CRPA. Purple line represents DALYs due to BSIs caused by PRSP. Circles represent medians and whiskers represent 95% uncertainty intervals.

We used the abridged life table for Japan from 2015 to 2018 (National Institute of Population and Social Security Research, 2020) to calculate the expected lifespan at the time of death in each BSI case and adopted the BSI outcome tree derived from the BCoDE project (European Centre for Disease Prevention and Control, 2019) because there is scarce evidence about the incidence of complications from BSIs. An outline of the outcome tree is shown in Supplementary Figure 1. The probability of each condition resulting from a complicated BSI and the respective disability weights are shown in Supplementary Table S2. As for uncomplicated cases, we do not take their acute phase burden into consideration when calculating DALYs. Although it is known that the length of stay in Japanese hospitals is generally much longer than in other developed countries (Tiessen et al., 2013), we have no precise data about length of stay due to bacteraemia in Japan. In addition, the burden of fatal cases and sequelae account for most of the DALYs due to bacteraemia. After considering these conditions, we excluded the burden of uncomplicated cases from the total disease burden.

We take into consideration the uncertainties inherent in this analysis by estimating uncertainty intervals (UIs). We drew 2,000 random samples for each of bed coverage, case fatality, probability, and utility of each health status according to their distribution (details are shown in Supplementary Table S1) and calculated the DALYs 2,000 times.

We set nine age groups: <1 year, 1-14 years, 15-24 years, 25-34 years, 35-44 years, 45-54 years, 55-64 years, 65-74 years, and ≥75 years. Both bed coverage and fatality/morbidity were taken into consideration to estimate 95% UIs. All statistical analyses were performed using R version 4.0.3 (R Core Team, 2018).

## Results

Figure 1 shows the estimated DALYs due to BSIs caused by the six major antimicrobial-resistant organisms examined in this study.

BSIs caused by MRSA, FQREC, and 3GREC account for most of the DALYs due to BSIs in Japan. Although the burden from MRSA decreased during the study period, that of FQREC increased.

Figure 2 shows a breakdown of DALYs according to causative organism and age group. Elderly people aged ≥65 years account for 62.2% of the total DALYs per 100,000 population (84.5/135.8) in 2018.

**Figure 2.**
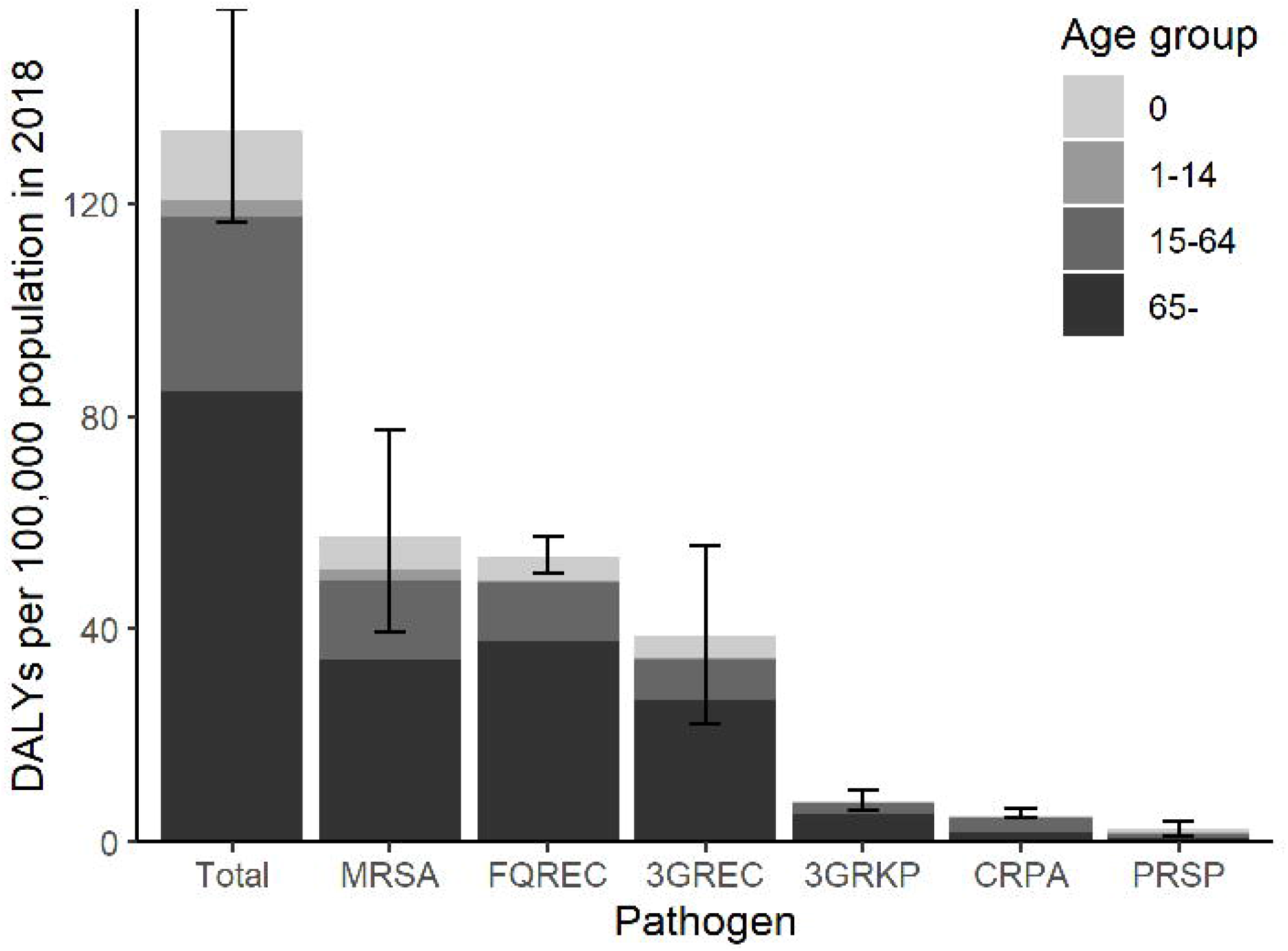
Breakdown of DALYs by age group in 2018. DALYs, disability-adjusted life years; BSIs, bloodstream infections; MRSA, methicillin-resistant *Staphylococcus aureus*; FQREC, fluoroquinolone-resistant *Escherichia coli*; 3GREC, third-generation cephalosporin-resistant *Escherichia coli*; 3GRKP, third-generation cephalosporin-resistant *Klebsiella pneumoniae*; CRPA, carbapenem-resistant *Pseudomonas aeruginosa*; PRSP, penicillin-resistant *Streptococcus pneumoniae* Lighter colours represent younger age groups. Whiskers represent 95% uncertainty intervals.

Table 1 shows the estimated number of death due to BSIs caused by each of the six major antimicrobial-resistant organisms from 2015 to 2018. MRSA, FQREC, and 3GREC accounted for 89.4% (8,433/9,447) of deaths due to BSIs caused by the six organisms in 2018. Figure 3 shows the proportions of YLLs and YLDs in total DALYs;

**Figure 3.**
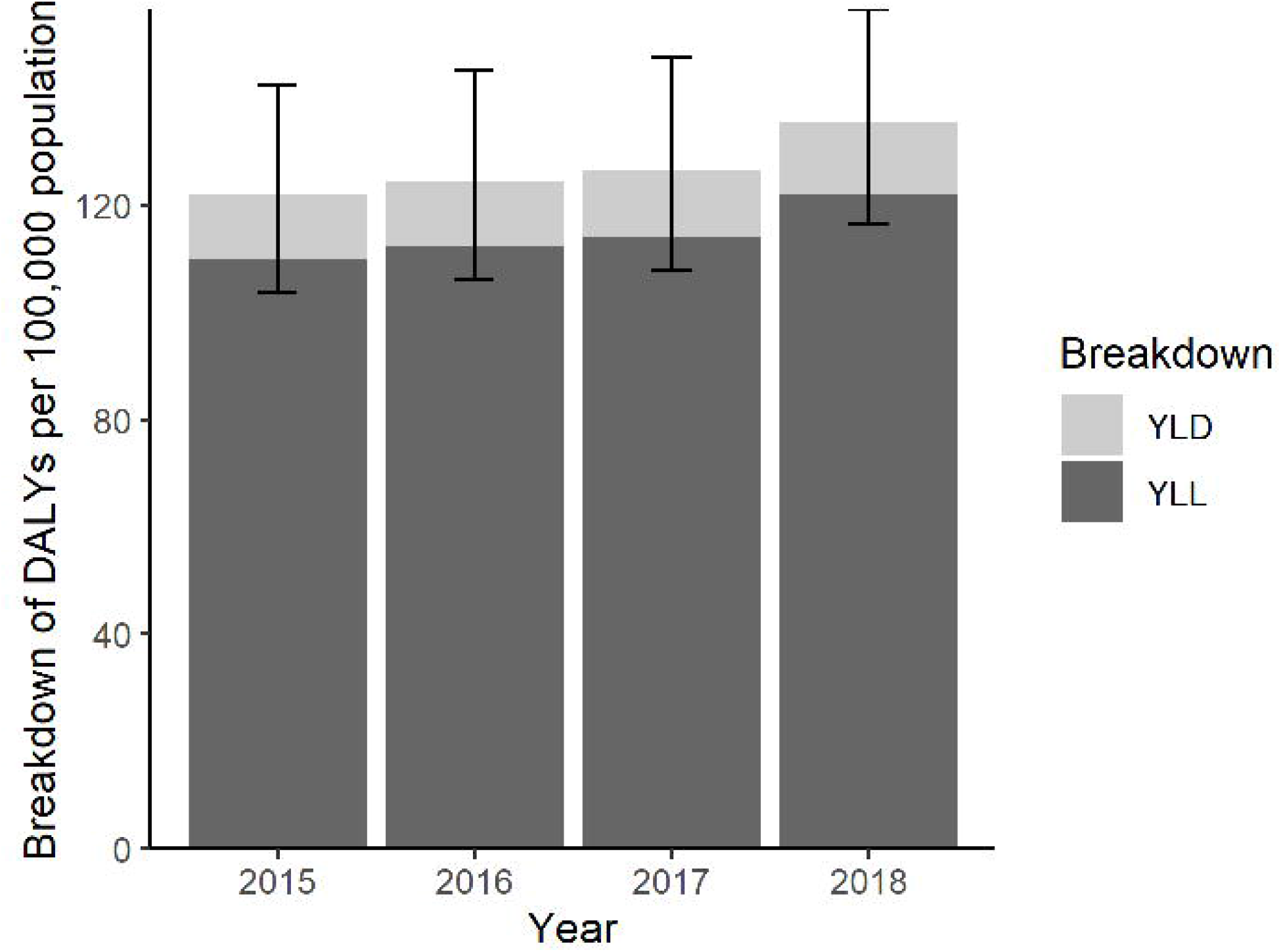
Breakdown of YLLs and YLDs among DALYs from 2015 to 2018. DALYs, disability-adjusted life years; YLLs, years of life lost; YLDs, years lived with disability Light grey bars represent the burden of YLDs. Dark grey bars represent the burden of YLLs. Whiskers represent uncertainty intervals.

YLLs accounted for 89.9% of DALYs (122.1/135.8) in 2018.

Figure 4 is a choropleth map of DALYs in each prefecture in 2018. The median value of DALYs by prefecture is 141.4 (IQR 106.5-167.7) per 100,000 population. Fukuoka Prefecture had the largest DALYs value (251.1/100,000 population) and Iwate prefecture had the smallest (57.0/100,000 population).

**Figure 4.**
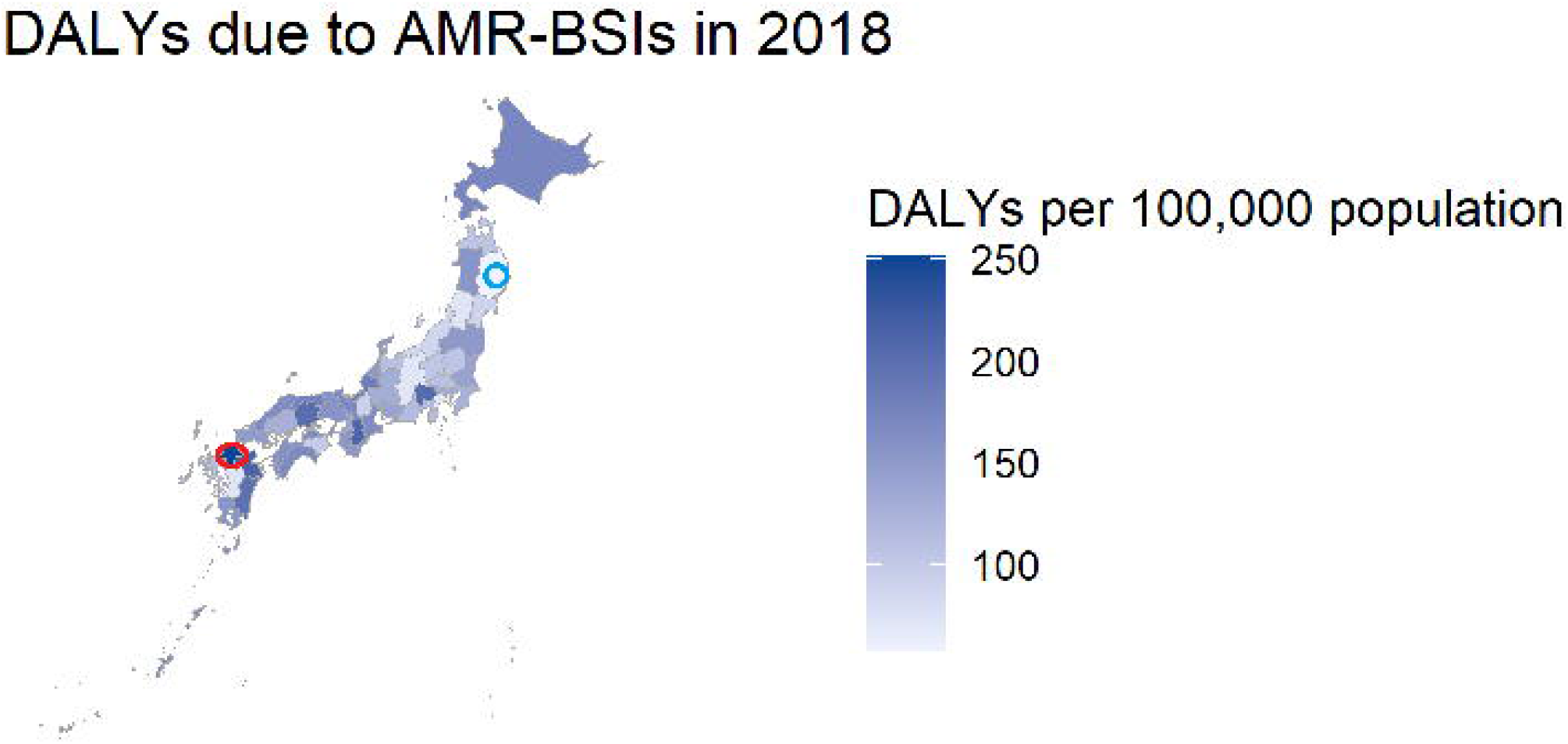
Choropleth map of DALYs due to AMR-BSIs by Japanese prefecture. DALYs, disability-adjusted life years; AMR-BSI, bloodstream infections caused by antimicrobial-resistant organisms Darker colours represent heavier burdens of AMR-BSIs. The red circle indicates the prefecture showing the highest value of DALYs (Fukuoka) and the blue circle indicates the prefecture showing the lowest value of DALYs (Iwate)

## Discussion

To our knowledge, this is the first study to quantitatively estimate the disease burden of BSIs due to antimicrobial-resistant organisms in the Western Pacific region. By comparing our results with those of the earlier study conducted in the EU/EEA area (Cassini et al., 2018), a more precise understanding of the disease burden caused by AMR can be obtained compared with evaluations based on single indicators such as number of deaths. In the present study, DALYs due to BSIs caused by the six major antimicrobial-resistant organisms was 135.8 per 100,000 population. This number is higher than that in the EU/EEA study (about 122 per 100,000 population), which included other minor organisms. A major factor contributing to the observed difference is the disease burden caused by MRSA, which is substantially higher in Japan than in the EU/EEA area (57.8 vs 20.9 per 100,000 population). Another factor might be the age distribution of the Japanese population, which has a higher old-age dependency ratio (47% in Japan and 31% in the EU/EEA in 2019) (National Institute of Population and Social Security Research, 2020; Eurostat, 2020). As suggested in Figure 2, the burden derived from elderly patients aged ≥65 years might be considerably larger in Japan than in EU/EEA countries. In contrast, the disease burden of 3GRKP in Japan is lower than that in the EU/EEA (7.8 vs 22.5 per 100,000 population).

It is noteworthy that Cassini and colleagues did not discount their results, and thus we should consider the possibility of overestimation by the previous study (Cassini et al., 2018). To more precisely compare the results of our study with theirs, we calculated DALYs without discounting, and the total DALYs caused by the six major antimicrobial-resistant BSIs in Japan was 162.4 (95%CI: 137.7-189.7) per 100,000 population in 2015. As discussed in the previous paragraph, a major factor contributing to this difference is the burden of MRSA because DALYs due to MRSA BSIs in 2015 in Japan was 75.6 (51.6-101.3) without adjustment, whereas that in EU/EEA countries was 20.9 (19.0-22.7). Although 3GREC also showed higher DALYs in Japan (40.0, 95%UI: 23.1-57.6) compared with EU/EEA countries (29.9, 95%UI: 26.4-33.6), the difference between the two areas was smaller than that for MRSA. Cassini and colleagues used the life table developed by Murray and colleagues (Murray et al., 2012), and thus the difference in life expectancy should be trivial; indeed, it did not have a substantial impact on this comparison. Considering the results of these comparisons, the AMR disease burden in Japan might be higher than that in EU/EEA countries, and MRSA can be considered an important target organism for AMR countermeasures in Japan in addition to *E. coli*. One of the strengths of the present study is that our results are also useful for understanding chronological trends within the same country. As shown in Figures 2 and 4, MRSA and drug-resistant *E. coli* are the largest causes of disease burden due to BSIs among antimicrobial-resistant bacteria in Japan during the study period. Interestingly, the MRSA burden has gradually decreased each year, whereas that of FQREC has increased. This trend is in line with the findings of our previous study (Tsuzuki et al., 2020) as well as studies conducted in other countries (Gagliotti et al., 2011; Musicha et al., 2017; Perencevich and Diekema, 2010; Wilson et al., 2011). These findings suggest that MRSA and FQREC would be appropriate target organisms for countermeasures against AMR. At the same time, the findings suggest that the importance of countermeasures against gram-negative bacilli will grow.

Additionally, our findings might be useful for other countries in the Western Pacific region. Given that AMR has become a major public health concern in this part of the world (Adamson et al., 2020; Argimón et al., 2020; Han and Zhang, 2020; Opatowski et al., 2020), our findings might be useful for comparing each country’s situation, although direct comparisons might be difficult due to differences in healthcare systems and other aspects.

The present study has several limitations. First, we borrow many of the parameters required for estimating DALYs from previous studies. For instance, the probability of having sequelae in each BSI case was derived from Cassini et al. and implemented using the BCoDE toolkit. We conducted elaborate simulations to reflect uncertainties related to the data; however, fatality and morbidity values should differ according to the country. It is more desirable to conduct similar estimations based on epidemiological data specific to Japan; therefore, establishing systems that enable us to obtain such information will be a future challenge. Second, we did not include the burden of BSIs themselves in the current analysis, mainly due to scarcity of data about length of stay in Japanese hospitals. Consequently, we excluded the burden of uncomplicated cases in its entirety. Although BSI is basically an acute curable disease, YLLs and YLDs should explain almost all of its disease burden; however, this might lead to underestimation. Third, we used the same parameter values for fatality and morbidity in all years. In addition, we excluded a small number of patients whose ages were not known (because it is not a mandatory input item in the national surveillance); therefore, this might be another cause of underestimation. It is possible that fatality was lower in 2018 than in 2015, which might be a cause of overestimation.

In conclusion, although the results should be interpreted cautiously in consideration of their limitations, the present study provides a quantitative estimation of AMR disease burden in Japan. The results should be useful for comparisons with studies conducted in other countries as well as the past situation within the same country. Nevertheless, further efforts are needed to resolve the limitations and obtain more precise results.

## Data Availability

The data used in this study will be made available by the corresponding author upon reasonable request.

## Contributors

ST and NO conceived the study. ST constructed the model, ran the simulations, and drafted the first manuscript. KY and KS aggregated and managed the raw data. KY, NM, KS, MS, and NO critically reviewed the manuscript. All authors approved the final version of the manuscript.

## Acknowledgments

We thank Toshiki Kajihara and Aki Hirabayashi for helpful discussions.

