## Supplementary file for "Disease burden of bloodstream infections caused by antimicrobial-resistant bacteria: a population-level study—Japan, 2015-2018"

**Supplementary Figure S1. Disease outcome tree of bloodstream infection**

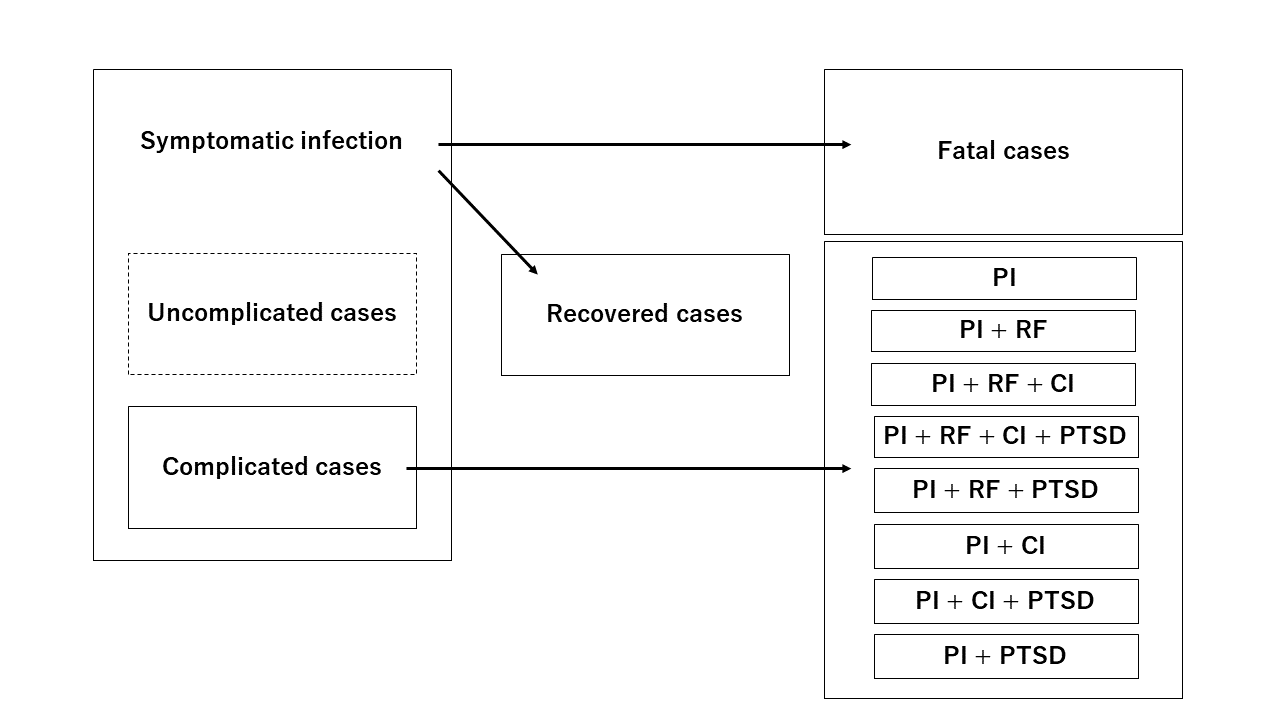

PI, physical impairment; RF, renal failure; CI, cognitive impairment; PTSD, post-traumatic stress disorder

**Supplementary Table S1. Fatality of blood stream infections by each causative organism**

|  | **Fatality** | **Reference** |
| --- | --- | --- |
| **Methicillin-resistant *Staphylococcus aureus*** | **0.25** | (Takeshita et al., 2017) |
| **Fluoroquinolone-resistant *Escherichia coli*** | **0.22** | (Abernethy et al., 2015) |
| **Third-generation cephalosporin-resistant *E. coli**** | **0.21** | (Nagao, 2013) |
| **Third-generation cephalosporin-resistant *Klebsiella pneumoniae*** | **0.32** | (Gallagher et al., 2014) |
| **Carbapenem-resistant *Pseudomonas aeruginosa*** | **0.35** | (Cassini et al., 2018) |
| **Penicillin-resistant *Streptococcus pneumoniae*** | **0.16** | (Takeshita et al., 2017) |

**Supplementary Table S2. Probability and disability weight of each health status**

|  | **Probability** | | **Disability weight** | |
| --- | --- | --- | --- | --- |
|  | **Mode** | **Range** | **Mode** | **Range** |
| **PI** | **0.584** | **0.441-0.734** | **0.033** | **0.012-0.052** |
| **PI + RF** | **0.288** | **0.071-0.489** | **0.288** | **0.071-0.489** |
| **PI + RF + CI** | **0.0026** | **0.001-0.0044** | **0.328** | **0.117-0.516** |
| **PI + RF + CI + PTSD** | **0.0005** | **0.0002-0.001** | **0.388** | **0.194-0.56** |
| **PI + RF + PTSD** | **0.0013** | **0.0008-0.002** | **0.353** | **0.154-0.534** |
| **PI + CI** | **0.236** | **0.0958-0.378** | **0.084** | **0.058-0.107** |
| **PI + CI + PTSD** | **0.0483** | **0.0187-0.0855** | **0.166** | **0.136-0.194** |
| **PI + PTSD** | **0.118** | **0.0799-0.173** | **0.119** | **0.09-0.146** |

PI, physical impairment; RF, renal failure; CI, cognitive impairment; PTSD, post-traumatic stress disorder

We assume that all of these follow a PERT distribution in accordance with the reference.

All values are derived from the ECDC BCoDE toolkit, version 2.0.0.
